# Identification and Genomic Assessment of Daptomycin-, Linezolid-, Vancomycin-resistant *Enterococcus faecium* During Protracted Infection

**DOI:** 10.1101/2022.03.28.22272907

**Authors:** Nathan B. Pincus, Tejas Joshi, Samuel W.M. Gatesy, Omar Al-Heeti, W. Justin Moore, Kelly E. R. Bachta

## Abstract

Linezolid and daptomycin resistance among *Enterococccus faecium* (*Efm)* isolates, while rare, is a major challenge for clinicians who are often limited to broad-spectrum or combination antibiotic therapies for management. Combination therapy with a beta-lactam has been reported, but limited clinical evidence exists to support its use. We describe the clinical management of a prolonged *Efm* intraabdominal (IA) infection and subsequent bacteremia, along with observed multidrug resistance development and use of serial whole genome sequencing to better understand resistance mechanisms. Combination antimicrobial therapy with daptomycin (DAP) and ceftaroline (CPT) was used to treat the patient’s catheter-associated daptomycin-, linezolid-, vancomycin-resistant *Efm* (DLVRE) bloodstream infection. *In vitro* antimicrobial testing of this DLVRE revealed only minor synergy between daptomycin and ceftaroline; However, the patient’s bacteremia cleared following initiation of combination therapy in conjunction with catheter removal. Sequencing of the patient’s DLVRE revealed multiple genomic mutations which explain both linezolid and daptomycin resistance and the presence of a plasmid containing known resistance determinants for vancomycin. Daptomycin resistance was attributed to the presence of chromosomal mutations in *liaS* (Thr120Ala), *liaR* (Trp73Cys), and *cls* (Asp13Ile), while linezolid resistance was attributed to the presence of the G2576T variant allele in some of 23S rRNA gene copies. Sequential whole genome sequencing of two additional bacterial isolates from the same patient revealed protracted colonization with a single DLVRE clone and suggested the development of bacterial subpopulations. Pairing clinical isolate susceptibilities with whole genome sequencing should be encouraged in clinical practice to better inform antimicrobial management in cases of multidrug resistance.

## INTRODUCTION

Multidrug resistant *Enterococcus faecium* (*Efm*) is an important cause of nosocomial infections including bacteremia, intraabdominal (IA) and urinary tract infections (UTIs). The incidence of enterococcal infections has increased steadily since the late 1970s (1) and enterococci are second only to *Escherichia coli* as a cause of nosocomial infection (2). In disease states, enterococci are believed to escape from the gastrointestinal (GI) tract, where they are natural colonizers and an important part of the GI microbiota (3). As GI tract residents, enterococci are frequently exposed to serial courses of antibiotics and have a remarkable ability to persist in part due to the plasticity of their genome (4). Antimicrobial resistance (AMR) arises through both genomic mutation and acquisition of mobile elements such as transposons and plasmids (5). Most enterococci demonstrate low-level intrinsic resistance to beta lactam antibiotics, moderate resistance to aminoglycosides and high resistance to most cephalosporins and clindamycin (6). Natural ampicillin resistance in *Efm* is largely attributed to penicillin-binding protein (PBP5) encoded by the bacterial chromosome, with variation in degree of resistance due to variable gene expression (7, 8) and the presence of a low-affinity allele associated with increased levels of resistance in the dominant hospital-associated *Efm* clade (1, 9, 10). Vancomycin resistance is driven by replacement of the terminal D-alanine residue of peptidoglycan precursors with either D-lactate or D-serine (3, 11). These altered peptidoglycan fragments are generated through expression of multiple gene clusters (*van* operons) which can be present on either the chromosome or transferable mobile plasmids (11).

Treatment of VRE often requires the use of newer antimicrobial agents including linezolid (LZD) and daptomycin (DAP) (12, 13). Though overall resistance remains rare (<1% for linezolid and <2% for daptomycin) (14, 15), resistance to both agents emerged shortly after these antibiotics were clinically introduced and is rising steadily (12, 15-19). Specifically, linezolid resistance is mediated by both chromosomal mutations and gene acquisition. Resistance to linezolid is linked to mutations in 23S ribosomal RNA genes and acquisition of plasmid-borne genes *cfr* and *optrA* encoding a ribosomal methyltransferase and an ATP-binding cassette transporter, respectively (15). Enterococcal resistance to daptomycin is more common in *Efm* than *E. faecalis* (1, 14), and resistance has been linked to mutations in two major groups of genes. The first (*liaFSR* and *yycFGHIJ*) encodes regulatory pathways that coordinate the stress response of the bacterial cell envelope. The second encodes enzymes that metabolize phospholipids, including glycerophosphoryl diester phosphodiesterase (*gdpD*) and cardiolipin synthetase (*cls*) (20, 21). The understanding of both linezolid and daptomycin resistance in enterococci continues to evolve (15, 20, 21).

Although they represent <1% of all enterococcal infections, the prevalence of daptomycin-, linezolid-, and vancomycin-resistant *Efm* (DLVRE) is increasing (14, 15, 22). Risk factors associated with the development of DLVRE include neutropenia, invasive procedures and cumulative antibiotic exposure, notably exposure to daptomycin, within the past year (23, 24). There are limited data on efficacious treatments for such infections. Combination therapy of daptomycin and a beta lactam antibiotic is thought to act synergistically to sensitize the bacterial cell membrane and improve the penetration of daptomycin (25). This combination is being explored for treatment (26, 27). *In vitro* studies with VRE revealed that ceftaroline (CPT) increased the bactericidal activity of daptomycin and lowered the daptomycin minimal inhibitory concentration (MIC) when co-administered (27-30). However, data are limited on the impact of dual antimicrobial therapy on clinical outcomes of patients with DLVRE infections.

Here, we report successful treatment of DLVRE catheter-associated bloodstream infection (CLABSI) in the setting of protracted IA abscess. We applied whole genome sequencing to characterize the genetic changes underlying emerging linezolid and daptomycin resistance in this case of persistent infection.

## METHODS

### Antimicrobial Resistance Testing

Antimicrobial resistance testing was performed at Northwestern Memorial Hospital and Northwestern University using the Vitek2 platform (bioMerieux) and Epsilometer testing (E-test). Additional synergy testing was performed on Mueller Hinton II (MHII) agar using E-test strips (Lisofilchem s.r.l). Briefly, bacteria were grown overnight at 37°C on blood agar plates. A suspension at optical density 600 (OD 600) of 0.08 to 0.1 was created by resuspending bacterial colonies in phosphate buffered saline (PBS). This suspension was subsequently confluently spread across the surface of a MHII agar plate. Plates were dried for five minutes and then a single DAP E-test strip was placed in the center of the plate and incubated for ten minutes. This DAP E-test strip was then removed and saved. In the same location and orientation as the DAP E-test, a CPT E-test strip was placed and the original DAP E-test strip replaced directly on-top (31). DAP and CPT single E-test controls were laid side-by-side on the same plates. The plates were incubated at 37°C for 24 hours, and the minimal inhibitor concentrations (MIC) were interpreted.

### Whole genome sequencing and genome assembly

Genomic DNA from Isolate 3 was isolated from a single colony grown overnight in 5 mL of tryptic soy broth using the Promega Maxwell 16 instrument. A short-read sequencing library was prepared with a plexWell™ 96 kit (seqWell), and sequencing was performed on an Illumina MiSeq with a v3 reagent kit yielding 709,596 301 bp paired-end reads totaling 427 Mbp. Illumina reads were quality trimmed using Trimmomatic v0.36 (32), resulting in 450,275 paired-end reads totaling 179 Mbp of sequence for approximate genomic coverage of 60-fold. Long-read sequencing was performed on the MinION platform with a FLO-MIN106 flow cell after library preparation with an Oxford Nanopore SQK-LSK109 ligation sequencing kit. Base calling and read-quality filtering was performed using Guppy v3.4.5 with the R9.4.1 high-accuracy model, yielding 248,837 reads totaling 1,339 Mbp. Using Filtlong v0.2.0, Nanopore reads were filtered to retain the best 95% of sequence with a minimum read length of 1000 bp, yielding 196,162 reads totaling 1,272 Mbp of sequence for an approximate coverage of 424-fold. Hybrid genome assembly was performed based on trimmed Illumina reads and filtered nanopore reads using Unicycler v0.4.8 (33) with default settings. The resulting complete genome was annotated using the NCBI Prokaryotic Genome Annotation Pipeline v5.3 (34). *In silico* sequence typing was performed by comparing our existing genomic sequences to *Efm* allele sequences and MLST profiles obtained from the PubMLST database (35). Isolate 3 plasmid replicons were identified using PlasmidFinder v2.1 (36) and compared to previously-described plasmids by BLAST search against *Enterococcus* sequences in the NCBI nt database.

Genomic DNA preparation and short-read sequencing library preparation for Isolates 4 and 5 were performed as described above. Sequencing was performed on an Illumina NovaSeq instrument yielding 150 bp paired-end reads totaling 162 Mbp for Isolate 4 and 316 Mbp for Isolate 5. Read trimming was performed as described above yielding 118 Mbp of sequence for Isolate 4 for an approximate coverage of 39-fold and 233 Mbp of sequence for isolate 5 for an approximate coverage of 78-fold. Draft genomes of Isolate 4 and 5 were assembled using SPAdes v3.9.1 with careful and automatic read coverage cutoff options (37) and filtered to remove contigs less than 200 bp or 5-fold mean coverage. Draft genomes were annotated using the NCBI Prokaryotic Genome Annotation Pipeline v5.3 (34). *In silico* sequence typing was performed as described above.

### Antimicrobial resistance gene identification

Known single nucleotide variants (SNVs) and acquired genes associated with AMR were identified in assembled genomes using AMRFinderPlus v3.10.16 (38) and ResFinder v4.1 (39). Illumina reads were screened for the presence of linezolid resistance mechanisms, including 23S rRNA gene variations and acquired genes, using LRE-Finder v1.0 (40).

### Pairwise genome comparisons

Isolates 4 and 5 were aligned to the Isolate 3 complete genome and SNVs between strains were identified as previously described (41). Briefly, trimmed Illumina reads were aligned to the Isolate 3 chromosome and plasmids using BWA v0.7.15 (https://arxiv.org/abs/1303.3997v2) and the resulting alignments sorted and indexed using samtools v0.1.19-44428cd (42). SNV calling from these alignments was performed using bcftools_filter.pl (https://github.com/egonozer/bcftools_filter). To better understand the genetic context of missing AMR genes in the Isolate 4 draft genome, Isolate 4 and 5 draft genome contigs were aligned to plasmid 3 of Isolate 3 using BRIG v0.95 (43).

### Data availability

Sequencing and genome assemblies have been deposited to NCBI under BioProject accession number PRJNA787599, with Isolate 3 termed NMVRE-001 (SAMN23828484), Isolate 4 termed NMVRE-002 (SAMN23828912), and Isolate 5 termed NMVRE-003 (SAMN23828937). Reads are available under SRA accessions SRR17230443-SRR17230446. Assemblies are available under GenBank accessions GCA_021228615.1, GCA_021364775.1, and GCA_021364755.1.

### Bioethics Statement

This research study (STU00214816) was submitted for review to the Northwestern University Institutional Review Board and, because of the retrospective nature of this work, was deemed “research that does not involve human subjects”. Further IRB review and approval was, therefore, not required. In good faith, the case report was discussed openly with the patient in question and the patient verbally consented to its publication.

## RESULTS

### Case Presentation

A middle-aged patient with a history of small bowel obstruction requiring multiple laparoscopic abdominal surgeries presented to our institution for elective incisional hernia repair. The patient underwent lysis of adhesions and small bowel resection with primary anastomosis and mesh closure. The patient’s post-operative course was complicated by the development of loculated IA abscesses secondary to an enteric leak at the anastomotic site. A drain was placed into the largest abscess and aspiration cultures were obtained on post-operative day (POD) 10 revealing a polymicrobial abscess including *Escherichia coli, Enterococcus faecalis*, coagulase-negative *Staphylococcus, Streptococcus constellatus, Candida albicans* and vancomycin-resistant *Efm* (Table 1, Isolate 1, VRE). Given an allergy to penicillin, the patient was initially treated with aztreonam and metronidazole. When cultures returned, the patient was transitioned to culture-directed therapy (meropenem, fluconazole and linezolid) for four weeks (Figure 1). At the completion of this course, clinical and radiographic improvement was achieved.

**TABLE 1.** Minimal Inhibitory Concentrations (MICs µg/mL) for the patient’s isolates of *Efm*

| Antibiotic | Isolate 1 | Isolate 2 | Isolate 3<sup>b</sup> | Isolate 4<sup>b</sup> | Isolate 5<sup>b</sup> |
| --- | --- | --- | --- | --- | --- |
|  | (VRE) | (LVRE) | (DLVRE) | (DLVRE) | (DLVRE) |
| Ampicillin | ≥32 R | ≥32 R | ≥32 R | ≥32 R | ≥32 R |
| Daptomycin | -- | 2 SDD | 12 R<sup>a</sup> | 16 R<sup>a</sup> | 16 R<sup>a</sup> |
| Linezolid | 2 S | 128 R<sup>a</sup> | 48 R<sup>a</sup> | >256 R<sup>a</sup> | 12 R<sup>a</sup> |
| Vancomycin | ≥32 R | ≥32 R | ≥32 R | ≥32 R | ≥32 R |

R, resistant; S, susceptible; SDD, susceptible dose dependent; --, not reported.
<sup>a</sup>MICs determined by E-testing assays.
<sup>b</sup>Sequenced isolate.

**FIGURE 1.**
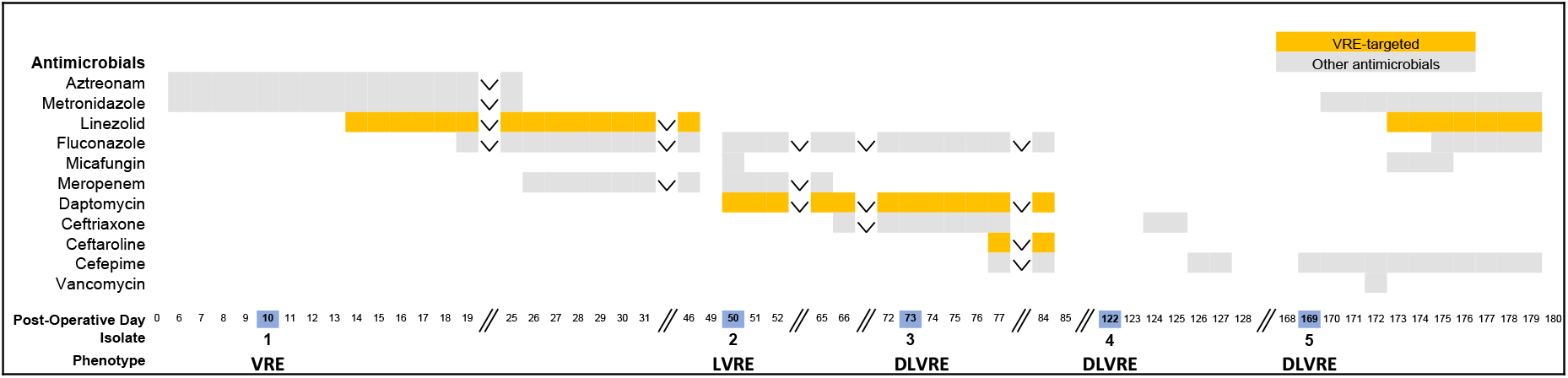
Timeline of antimicrobial treatment and *E. faecium* isolate recovery. Post-operative day and *Efm* isolate recovery profiles (V, vancomycin; L, linezolid, D, daptomycin; bottom) are shown together with antibiotic and antifungal treatment regimens. Vancomycin-resistant enterococci (VRE) targeted antimicrobials are highlighted in orange and other antimicrobials are presented in light grey. Isolate post-operative day recovery dates are highlighted in blue. Inverted carrots signify that the antimicrobial agent in question was continued throughout the time period in question. Two vertical forward slashes denote time breaks.

Four days after stopping antibiotics (POD 50), the patient developed recurrent abdominal pain and leukocytosis of 26.6K cells/µL. Meropenem, fluconazole and daptomycin were initiated. Daptomycin was chosen out of concern for long-term linezolid toxicity. Repeat IA cultures on POD 50 demonstrated a new linezolid resistant-VRE (Table 1, Isolate 2, LVRE). The patient was ultimately treated with intravenous ceftriaxone, fluconazole and daptomycin. On POD 73, following an episode of emesis directly onto an existing right brachial peripherally inserted central catheter (PICC), the patient developed a fever. Blood cultures from the periphery and PICC line both revealed new daptomycin resistant-LVRE (Table 1, Isolate 3, DLVRE). The PICC line was removed and blood cultures cleared the following day. Ceftaroline was added when the daptomycin non-susceptibility results became available. After one week of combination antimicrobial therapy, the patient had radiographic decrease in the size of the IA abscess, resolution of leukocytosis and reduction of fevers. The patient was discharged home on POD 86 off antibiotics.

Five weeks after discharge (POD 122), the patient was readmitted with left lower quadrant (LLQ) abdominal pain and increased drainage from around an abdominal drain site. CT scan revealed an additional abdominal wall fluid collection for which the patient underwent incision and drainage. Again, cultures were polymicrobial growing *Klebsiella oxytoca, Enterobacter cloacae, Streptococcus anginosus, E. coli, E. faecalis* and DLVRE (Table 1, Isolate 4). The patient was diagnosed with an enterocutaneous fistula and briefly treated with ceftriaxone followed by cefepime. In an effort to spare the patient prolonged courses of broad-spectrum antibiotics and in light of the patient’s clinical stability, the patient was discharged home off of all antibiotics on POD 128.

The patient was readmitted again on POD 169 following a fall. While hospitalized, the patient developed a new leukocytosis to 16.4K cells/µL, worsening LLQ abdominal tenderness and erythema. An abdominal CT scan demonstrated a new 5.8 cm abscess in the left anterior abdominal wall. The abscess was drained and cultures grew *Pseudomonas aeruginosa, K. oxytoca, E. cloacae, C. albicans* and DLVRE (Table 1, Isolate 5). The patient received a 10-day course of antimicrobials including cefepime, linezolid, metronidazole and fluconazole. Of note, Isolate 5 was initially reported as susceptible to linezolid (MIC of 2 µg/mL on VITEK2 platform); However, upon confirmatory testing, Isolate 5 was determined to be resistant to linezolid with an MIC of 12 µg/mL by E-test (Table 1, Isolate 5). Despite this discrepancy, the patient’s clinical status improved as erythema, abdominal tenderness and leukocytosis resolved. After one week of inpatient observation off antibiotics, the patient was discharged home on POD 187. A detailed course of antimicrobial therapy is outlined in Figure 1.

### In vitro synergy testing reveals little synergy between DPT and CPT

Given that the patient’s CLABSI was secondary to DLVRE (Isolate 3) leaving limited options for therapy, we chose to empirically start combination therapy with daptomycin and ceftaroline (25-27, 30). We sought to validate our antimicrobial choice *in vitro* by conducting synergy antimicrobial resistance testing (31). Confirmatory testing (E-test) in the laboratory revealed individual MICs to daptomycin of 16 μg/mL and ceftaroline of 32 μg/mL. Synergy testing in the experimental laboratory revealed only modest synergy, with the addition of ceftaroline lowering the daptomycin MIC from 16 to 12 μg/mL (Fig. 2). Addition of ceftaroline did not change the clinical interpretation of daptomycin resistance in Isolate 3 (44).

**FIGURE 2.**
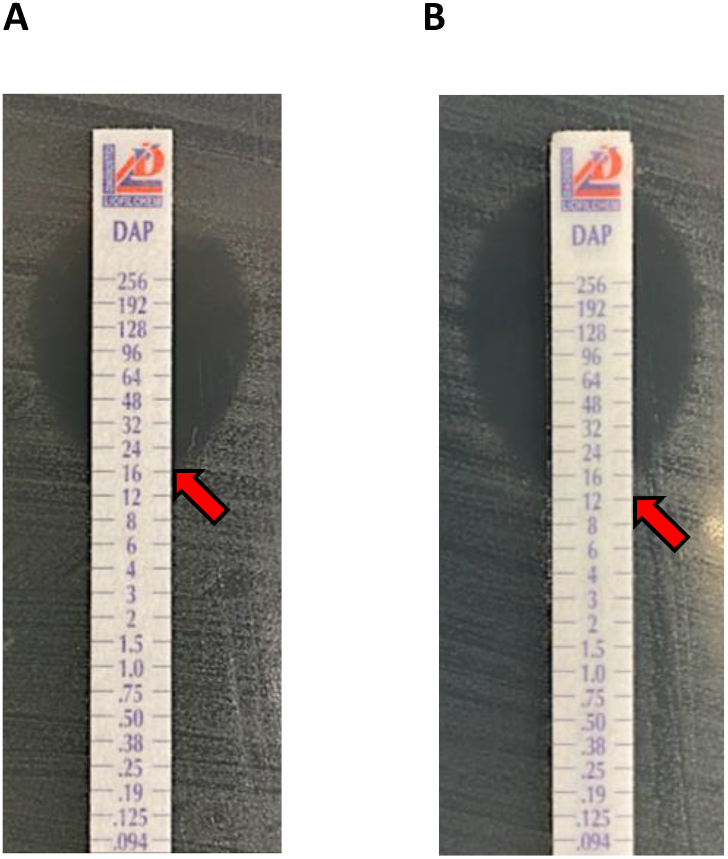
Synergy susceptibility testing of *Efm* Isolate 3. (A) DAP E-test of isolate 3 (16 μg/mL). (B) Combination E-testing of DAP and CPT of Isolate 3 (DAP MIC 12 μg/mL). Synergy was tested by first applying a DAP E-test to the plate, removing it, applying a CPT E-test on the exact same location, and placing the DAP E-test directly on top to read the susceptibility to DAP in the presence of CPT.

### Genomic evaluation of antimicrobial resistance in Isolate 3

We sought to use genomics to understand the mechanisms behind evolving drug-resistance in our DLVRE isolates. Unfortunately, we did not have access to Isolates 1 and 2 for comparison as they had been previously discarded. Whole genome sequencing of Isolate 3 confirmed the identity of a sequence type (ST) 584 *Efm*. ST584 is a member of the pandemic clonal complex 17 (CC17) that includes a collection of hospital-acquired *Efm* sequence types (45-47). The complete genome of Isolate 3 included a single chromosome of 2.8 Mbp and 5 plasmids of lengths 228.3 Kbp (p1), 50.1 Kbp (p2), 40.6 Kbp (p3), 11.9 Kbp (p4), and 1.9 Kbp (p5), respectively. The sequence present in these plasmids was similar to that previously described in *Efm* plasmids, although present in different configurations. For example, nearly all of the sequence present in plasmids 2 and 4 has been described in substantially larger, previously described plasmids (Supplementary Table S1) reaffirming that *Efm* plasmids are highly modular (48).

Examination of the complete genome of Isolate 3 for known antimicrobial resistance determinants reasonably explained the observed susceptibility pattern (Table 2). Isolate 3 contained the low-affinity penicillin-binding protein 5 allele (*pbp5-R*) found in the *Efm* nosocomial clade, clade A, that is associated with ampicillin resistance (1, 9, 10). On the 40.6kb p3, we identified the presence of a *vanA* operon (Figure 3), explaining the observed vancomycin resistance. The *vanA* operon was flanked by IS1216 family transposases, consistent with the known presence of *vanA* operons on plasmid-borne *Tn1546*-type transposons (49). Daptomycin resistance was attributed to the presence of chromosomal mutations in *liaS* (Thr120Ala), *liaR* (Trp73Cys) and *cls* (Asp13Ile). Other resistance mechanisms identified included fluoroquinolone resistance mutations, *gyrA* (Ser83Tyr) and *parC* (Ser80Arg), aminoglycoside resistance genes *aac(*6’)-I, *aph*(3’)-IIIa and *ant*(6)-Ia, trimethoprim resistance genes *dfrG* and *dfr*F and tetracycline resistance genes *tetL* and *tetM* (Table 2).

**TABLE 2.**
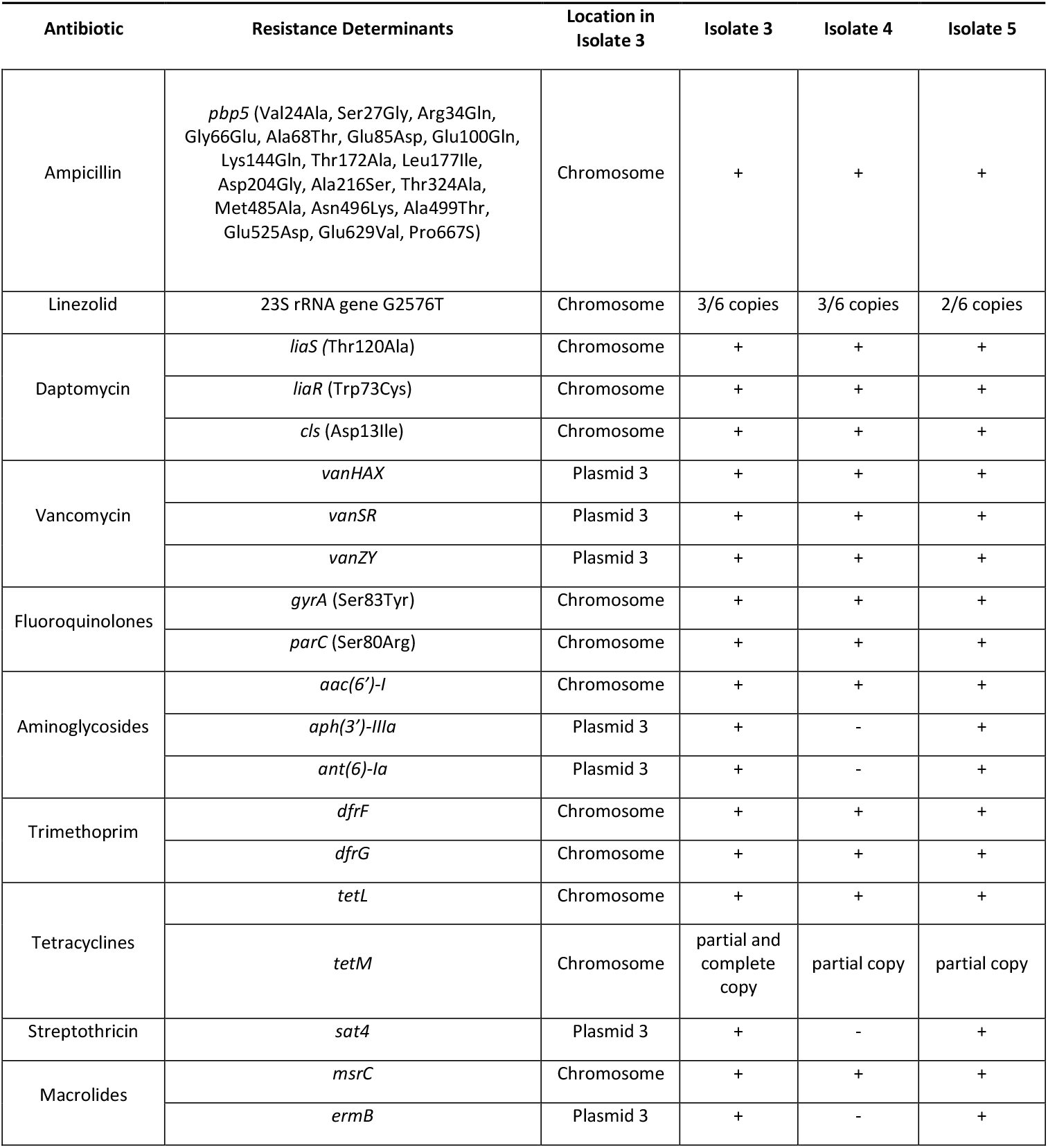
Genomic mechanisms of antimicrobial resistance identified in Isolates 3, 4, and 5.

| Antibiotic | Resistance Determinants | Location in Isolate 3 | Isolate 3 | Isolate 4 | Isolate 5 |
| --- | --- | --- | --- | --- | --- |
| Ampicillin | *pbp5* (Val24Ala, Ser27Gly, Arg34Gln, Gly66Glu, Ala68Thr, Glu85Asp, Glu100Gln, Lys144Gln, Thr172Ala, Leu177Ile, Asp204Gly, Ala216Ser, Thr324Ala, Met485Ala, Asn496Lys, Ala499Thr, Glu525Asp, Glu629Val, Pro667S) | Chromosome | + | + | + |
| Linezolid | 23S rRNA gene G2576T | Chromosome | 3/6 copies | 3/6 copies | 2/6 copies |
| Daptomycin | *liaS* (Thr120Ala) | Chromosome | + | + | + |
|  | *liaR* (Trp73Cys) | Chromosome | + | + | + |
|  | *cls* (Asp13Ile) | Chromosome | + | + | + |
| Vancomycin | *vanHAX* | Plasmid 3 | + | + | + |
|  | *vanSR* | Plasmid 3 | + | + | + |
|  | *vanZY* | Plasmid 3 | + | + | + |
| Fluoroquinolones | *gyrA* (Ser83Tyr) | Chromosome | + | + | + |
|  | *parC* (Ser80Arg) | Chromosome | + | + | + |
| Aminoglycosides | *aac(6')-I* | Chromosome | + | + | + |
|  | *aph(3')-IIIa* | Plasmid 3 | + | - | + |
|  | *ant(6)-Ia* | Plasmid 3 | + | - | + |
| Trimethoprim | *dfrF* | Chromosome | + | + | + |
|  | *dfrG* | Chromosome | + | + | + |
| Tetracyclines | *tetL* | Chromosome | + | + | + |
|  | *tetM* | Chromosome | partial and complete copy | partial copy | partial copy |
| Streptothricin | *sat4* | Plasmid 3 | + | - | + |
| Macrolides | *msrC* | Chromosome | + | + | + |
|  | *ermB* | Plasmid 3 | + | - | + |

**FIGURE 3.**
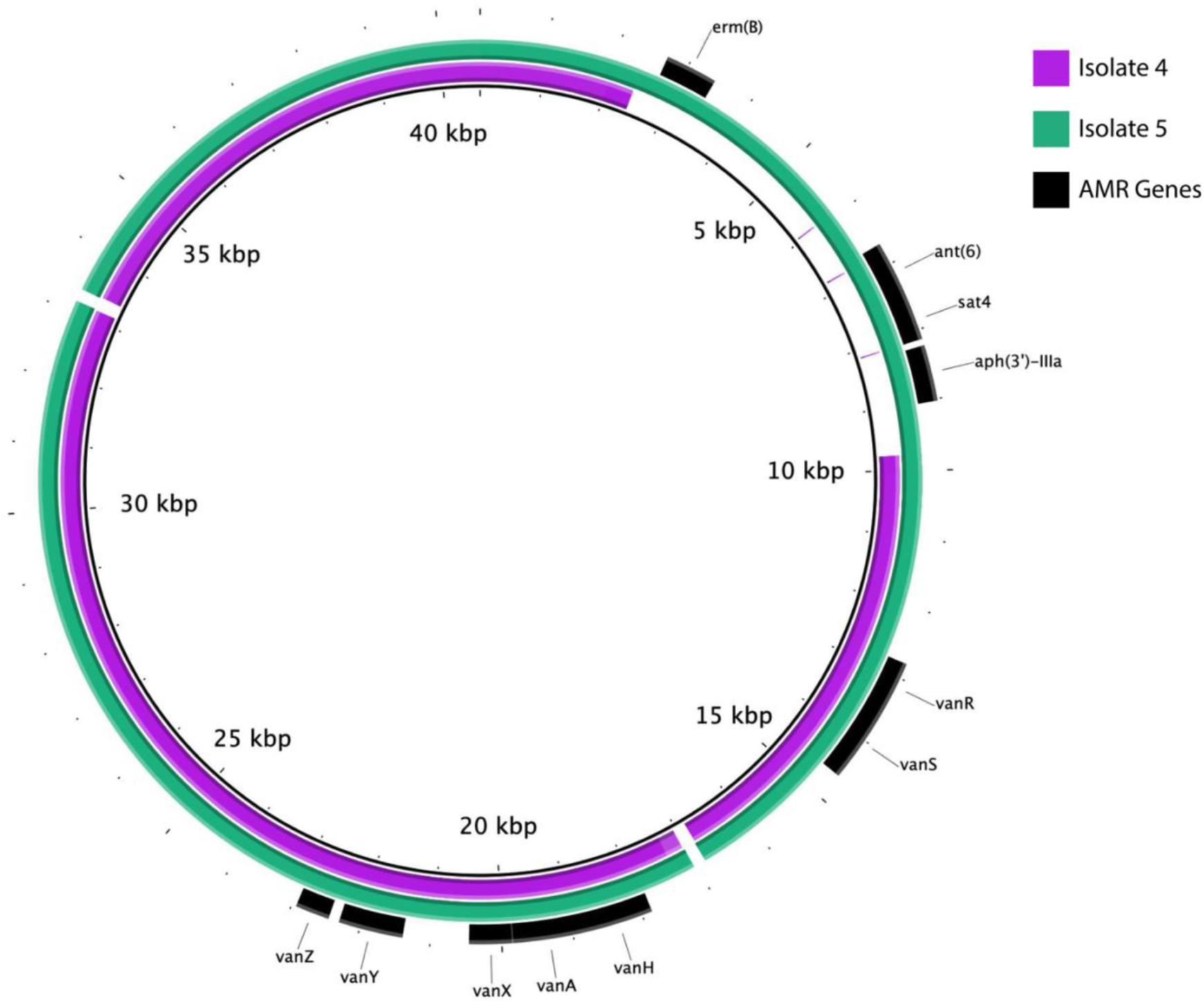
Alignment of plasmid 3 from Isolates 4 and 5 to the vancomycin resistance plasmid 3 of Isolate 3. Regions present in Isolate 4 and 5 draft genomes are indicated in purple and green respectively. Identified resistance genes are indicated in black.

The mechanism of linezolid resistance for Isolate 3 was not immediately apparent from interrogation of its complete genome sequence. However, a read-based analysis (40) estimated that Isolate 3 contained three copies of the variant G2576T allele in the six copy 23S rRNA gene (48% of aligned reads contained the G2576T variant allele). The absence of these variants in the complete genome was likely an artifact of the assembly method, which used nanopore reads to bridge short-read derived contigs (33), combined with the fact that they were present in only some copies of a multi-copy gene. Variant alleles in the 23S rRNA gene, particularly G2576T, are the most common mechanism of linezolid resistance in *Efm* (15). The level of linezolid resistance correlates with the number of variant alleles (40, 50). Other determinants associated with acquired linezolid resistance, including the 23S rRNA methyltransferase gene *cfr* and the ATP-binding cassette ABC-F gene *optrA*, were not present in the genome (Table 2).

### Evolution of *Enterococcus faecium* during protracted infection

Given the patient’s repeated admissions and repeated isolation of DLVRE, we chose to perform short-read sequencing on two additional *Efm* Isolates, Isolates 4 and 5. Isolates 4 and 5 were both clonal with Isolate 3 and belonged to the same sequence type, ST584. Isolate 4 had 2 chromosomal SNVs with respect to Isolate 3, while Isolate 5 had 13 chromosomal and 3 p1 SNVs (Supplementary Table S2). All SNVs present in Isolate 5 were previously observed in Isolate 4. While Isolate 4’s SNVs were located in intergenic regions, Isolate 5 also contained SNVs in predicted protein coding genes (Supplementary Table S3). Altogether, these findings strongly suggest persistent infection, or colonization with recurrent infection, by a specific DLVRE clone.

Isolates 4 and 5 contained the same elements associated with resistance to ampicillin, vancomycin, and daptomycin. Similar to Isolate 3, Isolate 4 contained an estimated three copies of the 23S rRNA gene containing the G2576T variant allele (56% G2576T), likely leading to observed linezolid resistance. Isolate 5, on the other hand, was estimated to encode two copies of the 23S rRNA gene containing the G2576T variant allele (33% G2576T) (Table 2). As previously stated (40, 50), copy number of the G2576T variant allele correlates with level of resistance, potentially explaining the lower linezolid MIC seen in Isolate 5 (Table 1). There may be other contributing factors given the substantially different MICs between Isolates 3 and 4 despite each possessing the same number of mutant alleles.

While the other 4 plasmids were fairly well conserved between Isolates 4 and 5 and Isolate 3, we noted the absence of a substantial portion of p3 in Isolate 4 (Figure 3, Supplementary Table S2), resulting in the loss of resistance genes *ermB, ant(6), sat4*, and *aph(3’)-IIIa* (Table 2, Figure 3). This deletion of approximately 7.4 Kbp was notably flanked by IS1216 family transposase genes, suggesting that it was located on a mobile element. Interestingly, this sequence was present in Isolate 5 (Figure 3). This implies that multiple subpopulations of this ST584 DLVRE clone developed during our case of protracted infection, as the intact p3 would need to be present on POD 122 when Isolate 4 was collected to be found again in subsequent Isolate 5 on POD 169.

## DISCUSSION

In this study, a patient with an IA abscess known to harbor VRE developed DLVRE bacteremia following prolonged sequential treatment with linezolid and daptomycin. Ultimately, the patient experienced an episode of CLABSI secondary to DLVRE. Limited data exist on appropriate antibiotic choice in such challenging clinical situations (16, 24). Based on prior clinical and experimental data (25-27, 29, 30), we elected to use daptomycin in conjunction with ceftaroline for synergy, and the patient achieved a desired clinical response: clearance of blood cultures and diminishing size of an IA abscess. To our knowledge, this is the first case report demonstrating a successful clinical outcome with the use of daptomycin and beta-lactam antimicrobial therapy for CLABSI associated with IA abscesses. In our case, it is difficult to delineate if successful treatment of bacteremia was achieved due to the removal of an infected central line, dual antimicrobial therapy or the combination of both. The generalizability of this combination approach is also limited by our experiences in a single patient. These results point to the need for further evaluation of daptomycin and beta-lactam antimicrobial combination therapy for DLVRE infections in larger clinical studies. This case also highlights the challenges of determining appropriate duration of antibiotics when a polymicrobial abscess persists. Our patient ultimately received a total of 57 days of broad-spectrum antibiotics. Repeat cultures revealed the development of progressive antimicrobial resistance over time and, despite our best stewardship efforts, the patient’s symptoms recurred. This outcome underscores the challenges that clinicians face when deploying culture-directed therapy in patients with protracted infectious reservoirs. In settings where complete source control is not achievable, there seems to be no clear answer on antibiotic choice or duration.

With the rise in VRE infections, clinicians are increasingly reliant on reserved antibiotic therapies such as daptomycin and linezolid as the backbone of successful treatment. By deploying whole-genome sequencing in a series of DLVRE isolates separated by 13 weeks, we were able to determine genetic explanations for the observed vancomycin, daptomycin and linezolid resistant phenotypes in our isolates. Our work, in conjunction with others, demonstrates the utility of whole-genome sequencing longitudinal samples during prolonged infection to map genomic changes responsible for acquired AMR (17, 22). Given the observation that there was plasmid heterogeneity in p3 between Isolates 3 and 5 and Isolate 4, our also work suggests that there are likely subpopulations of *Efm* that develop during protracted infection. Plasmid 3 was virtually identical in Isolates 3 and 5, which are separated by 96 days, but was missing a substantial segment in Isolate 4 which was isolated temporally between Isolates 3 and 5. The idea of a single species bacterial “cloud of diversity” in prolonged infection is not new, but the impact of this diversity on successful antimicrobial treatment and infection outcome is clinically underappreciated (4, 51, 52). We have shown that subpopulations may possess different armamentariums of AMR genes and vary in degree of resistance. These observations are missed by traditional methods of AMR detection, making targeted treatment decisions based on such methods problematic. Phenotypic resistance analysis from an isolated colony may mask small subpopulations that harbor more difficult-to-treat antimicrobial resistant phenotypes and lead to suboptimal antimicrobial therapy decisions.

With the increasing rates of MDR *Efm* infections, it is critical that we understand how *Efm* adapts to prolonged antimicrobial pressure and recognize that traditional clinical resistance testing may not mirror population level AMR phenotypes. Further work is needed to elucidate the best course of treatment for patients with VRE requiring long-term antibiotic therapy and for those who develop MDR *Efm* while on therapy (24). Synergy testing is currently not available in most clinical laboratories. Our study highlights the urgent need for rapid implementation of investigational MIC testing paired with genomic analysis to successfully support real-time clinical decision making.

## Data Availability

All data produced in the present study are contained in the manuscript. Genomic sequences are available at the accession numbers listed in the methods.

## ACKNOWLEDGMENTS

This work was supported by grants from the National Institutes of Health (NIH)/National Institute of General Medical Sciences (NIGMS) (T32 GM008152 awarded to N.B.P), an American Cancer Society (ACS) Clinician Scientist Development Grant (#134251-CSDG-20-053-01-MPC, awarded to K.E.R.B.), and a Northwestern University Emerging and Re-emerging Pathogen Program (EREPP) grant (awarded to K.E.R.B.). This work was also supported by the Northwestern Memorial Clinical Microbiology laboratory and the NUSeq Core Facility which is generously supported by the NCI CCSG P30 CA060553 award to the Robert H. Lurie Comprehensive Cancer Center. This work was supported in part through the computational resources and staff contributions provided by the Genomics Compute Cluster which is jointly supported by the Feinberg School of Medicine, the Center for Genetic Medicine, and Feinberg’s Department of Biochemistry and Molecular Genetics, the Office of the Provost, the Office for Research, and Northwestern Information Technology. The Genomics Compute Cluster is part of Quest, Northwestern University’s high-performance computing facility, with the purpose to advance research in genomics. The funders had no role in study design, data collection and analysis, decision to publish, or preparation of the manuscript. We would like to thank members of the Hauser and Ozer laboratories for their valuable comments during numerous discussions of this work.

## FIGURES and TABLES

**SUPPLEMENTARY TABLE S1.** Characterization of Isolate 3 plasmids.

| Plasmid | Length (bp) | Replicon | Representative Similar Plasmid<sup>a</sup> |  |  |  |  |
| --- | --- | --- | --- | --- | --- | --- | --- |
|  |  |  | GenBank Accession | Length | Query Coverage (%) | Query Identity (%) | PMID Reference |
| 1 | 228311 | repUS15 | CP064425.1 | 203164 | 80 | 99.96 | 33332354 |
| 2 | 59066 | unknown | CP012468.1 | 84196 | 92 | 100 | 28301471 |
| 3 | 40605 | rep17 | CP076330.1 | 40559 | 95 | 99.87 | 34747439 |
| 4 | 11983 | rep29 | CP065776.1 | 27166 | 99 | 99.16 | 33895846 |
| 5 | 1928 | rep14a (partial) | CP064442.1 | 1928 | 100 | 100 | 33332354 |

<sup>a</sup>Based on BLAST search against *Enterococcus* sequences in the NCBI nt database

**SUPPLEMENTARY TABLE S2.**
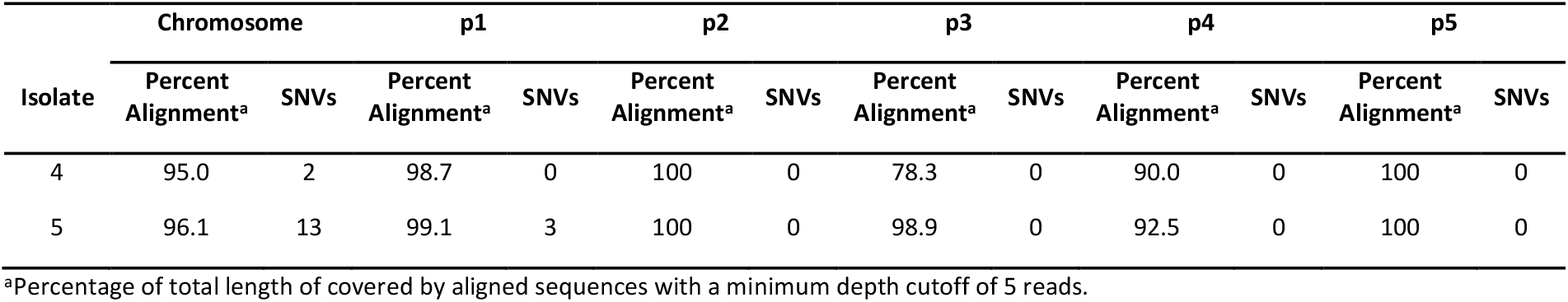
Alignment of Isolate 4 and 5 to the Isolate 3 Complete Genome.

**SUPPLEMENTARY TABLE S3.** SNVs Detected in Alignment of Isolate 4 and 5 to the Isolate 3 Complete Genome.

| Location | Site | Annotation in Isolate 3 | Isolate 4 | Isolate 5 |
| --- | --- | --- | --- | --- |
| Chromosome | 89200 | intergenic | - | + |
| Chromosome | 232118 | cation-translocating P-type ATPase | - | + |
| Chromosome | 413477 | intergenic | + | + |
| Chromosome | 1016951 | cardiolipin synthase | - | + |
| Chromosome | 1125507 | NAD(P)/FAD-dependent oxidoreductase | - | + |
| Chromosome | 1208228 | *rsmD* | - | + |
| Chromosome | 1263527 | branched-chain amino acid aminotransferase | - | + |
| Chromosome | 1342358 | ABC transporter substrate-binding protein | - | + |
| Chromosome | 1887988 | intergenic | + | + |
| Chromosome | 1896892 | Xaa-Pro peptidase family protein | - | + |
| Chromosome | 2107572 | alpha/beta fold hydrolase | - | + |
| Chromosome | 2547067 | intergenic | - | + |
| Chromosome | 2720433 | pseudogene | - | + |
| p1 | 167 | *repA* | - | + |
| p1 | 120818 | HAMP domain-containing histidine kinase | - | + |
| p1 | 177993 | Heavy metal translocating P-type ATPase | - | + |

